# Valence-dependent self-agency is disturbed in depression and anxiety

**DOI:** 10.1101/2022.07.11.22277423

**Authors:** Marishka M Mehta, Soojung Na, Xiaosi Gu, James W Murrough, Laurel S Morris

**Affiliations:** Department of Psychiatry, Icahn School of Medicine at Mount Sinai; Depression and Anxiety Center for Discovery and Treatment, Icahn School of Medicine at Mount Sinai; Center for Computational Psychiatry, Icahn School of Medicine at Mount Sinai

## Abstract

**Background:** The sense of agency, or the belief in action causality, is an elusive construct that impacts day-to-day experience and decision-making. Despite its relevance in a range of neuropsychiatric disorders, it is widely under-studied and remains difficult to measure objectively in patient populations. We developed and tested a novel cognitive measure of valence-modulated agency perception in an in-person and online cohort.

**Methods:** The in-person cohort consisted of 52 healthy control subjects and 20 subjects with depression and anxiety disorders (DA), including major depressive disorder and generalized anxiety disorder. The online sample consisted of 254 participants. The task consisted of an effort task for monetary rewards with computerized visual feedback interference and trial-by-trial ratings of self versus other agency.

**Results:** All subjects across both cohorts demonstrated higher self-agency after receiving positive-win feedback, compared to negative-loss feedback when the level of computer inference was kept constant. Patients with DA showed reduced positive valence-dependent agency compared to healthy controls. Finally, in both cohorts, lower self-agency following negative-loss feedback was associated with worse anhedonia symptoms.

**Conclusion:** Together this work suggests how positive and negative environmental information impacts the sense of self-agency in healthy subjects, and how it is perturbed in patients with depression and anxiety.

## Introduction

The sense of agency, belief of action ownership, or belief in a direct relationship between action and outcome, can influence decision-making and motivated behavior. A higher sense of self-agency, also known as internal locus of control, describes a stronger link between internal self-agency and environmental outcomes, whereas a lower sense of self-agency, known also as external locus of control, describes outcomes that are not tied to ones’ actions. The sense of self-agency is a relatively stable trait (over 9 years), increasing slightly with age (Hovenkamp-Hermelink et al., 2019). Higher self-agency has been associated with resilience to stress in the form of lower cortisol response to stress induction (Bollini et al., 2004), greater incentive motivation (Declerck et al., 2006), greater exploratory behavior (Kayser et al., 2015), more flexible and goal-directed behaviors (Declerck et al., 2006) and overall well-being (DeNeve & Cooper, 1998). Higher self-agency is also associated with higher plasma dopamine metabolite levels (De Brabander & Declerck, 2004), and larger volumes of dopamine-rich fronto-striatal regions associated with cognitive and emotional control (i.e. ACC, striatum, anterior insula (Hashimoto et al., 2015). On the other hand, lower self-agency has been associated with negative emotionality (Yoshie & Haggard, 2013), learned helplessness (Soral et al., 2021), higher risk of negative symptoms related to apathy and anhedonia (Thompson et al., 2013) with early life experiences of low self-agency creating higher risk for later-life problematic anxiety (Chorpita & Barlow, 1998).

Given these relationships between self-agency and prominent behavioral disturbances known to be disturbed in depression and anxiety disorders, it is surprising that little empirical evidence measures this trait in patient groups. Limited evidence mainly from self-reported measures of self-agency in the form of general locus of control (internal versus external) points towards a role for disrupted self-agency in patients with anxiety disorders and depression. One study in patients with major depressive disorder (MDD) showed an inverse relationship between self-agency (Rotter’s LOC scale) and self-reported depressive symptoms (BDI) (Abdolmanafi et al., 2011). Another study in a large longitudinal cohort demonstrated that lower self-agency scores (CNSIE scale) at age 16 was associated with worse depressive symptoms at age 18, the former driven by worse early socioeconomic status (Culpin et al., 2015). Another longitudinal study in a large sample of subjects with a depressive or anxiety disorder found lower self-agency was associated with worse depression (Inventory of depression) and anxiety (BAI), with higher depression and negative life events predicting lower self-agency (Hovenkamp-Hermelink et al., 2019).

In this study, we propose a novel objective cognitive assessment of self-agency. We developed and operationalized an objective non-learning measure of self-agency in changing positive and negative environmental contexts and outcomes, to examine putative trait-level constructs of valence-dependent self-agency across groups. This task was tested in two samples, in the context of a laboratory-based in-person experiment as well as a larger online experiment. We predicted that patients with depression and anxiety disorders, or those with worse depressive symptoms, would have overall lower self-agency across both positive and negative contexts.

## Materials and Methods

### Experiment 1: In Person Participants

A community sample of volunteers between the ages of 18-65 were recruited through the Depression and Anxiety Center at the Icahn School of Medicine at Mount Sinai, New York. Subjects were included if they met criteria for MDD or GAD as their primary psychiatric diagnosis as determined by the Structured Clinical Interview for DSM-V Axis Disorders (SCID-V) conducted by a trained rater. Subjects with MDD or GAD were allowed comorbid mood or anxiety disorders, which are commonly comorbid (T. A. Brown et al., 2001). Healthy subjects were free from any current or lifetime psychiatric disorder. For all groups, subjects were excluded if they had an unstable medical illness, history of neurological disease, neurodevelopmental or neurocognitive disorder, or positive urine toxicology test. Subjects completed cognitive testing and completed self-reported scales on the same day designed to capture dimensional measure of anhedonia (Temporal Experience of Pleasure Scale, TEPS). All methods were performed in accordance with the relevant guidelines and regulations set by the Program for Protection of Human Subjects (PPHS)/Institutional Review Board (IRB) at Icahn School of Medicine at Mount Sinai–approved written informed consent and subjects were compensated for their time.

### Experiment 2: Online Participants

A sample of US-based volunteers between the ages of 18-45 were recruited through the Prolific web-based research platform (https://www.prolific.co/). A total of 318 participants were enrolled in the study. Subjects completed an online version of the cognitive task and self-reported scales in the same session. Attention checks were embedded throughout the self-report questionnaires and any subjects that failed more than one were excluded. Participants were also excluded if cognitive task performance-based data quality thresholds were not met. Performance-based data quality thresholds included performance of at least 10 button presses on average throughout the entire task and an overall average minimum reaction time of 1s, to exclude automatic responses without task engagement. The testing and data collection protocol was approved by the Program for Protection of Human Subjects (PPHS)/Institutional Review Board (IRB) at Icahn School of Medicine at Mount Sinai and subjects were compensated for their time.

### The Self-Agency Task (SAT)

The self-agency task was a 20-minute cognitive task programmed in *PsychoPy*. Throughout the task, subjects performed a simple effort task for reward over 108 trials (Figure 1). Subjects were instructed to press the left and right keyboard buttons quickly to move a visual bar on the computer screen to a target position. During baseline practice trials, reaction times were recorded and used to determine the subsequent threshold (x1.25). Each trial indicated the target bar position and context information on whether the trial was a win trial (for $0.50) or a loss trial (-$0.50). Subjects then performed the button presses to move the bar up to the target position as quickly as possible on each trial. Subjects received outcome feedback based on their performance (win trials: win money or nothing; loss trials: lose money or nothing). Performance was defined as successfully moving the bar to the target position in the available time, i.e. 125% of their average baseline reaction time.

**Figure 1.**
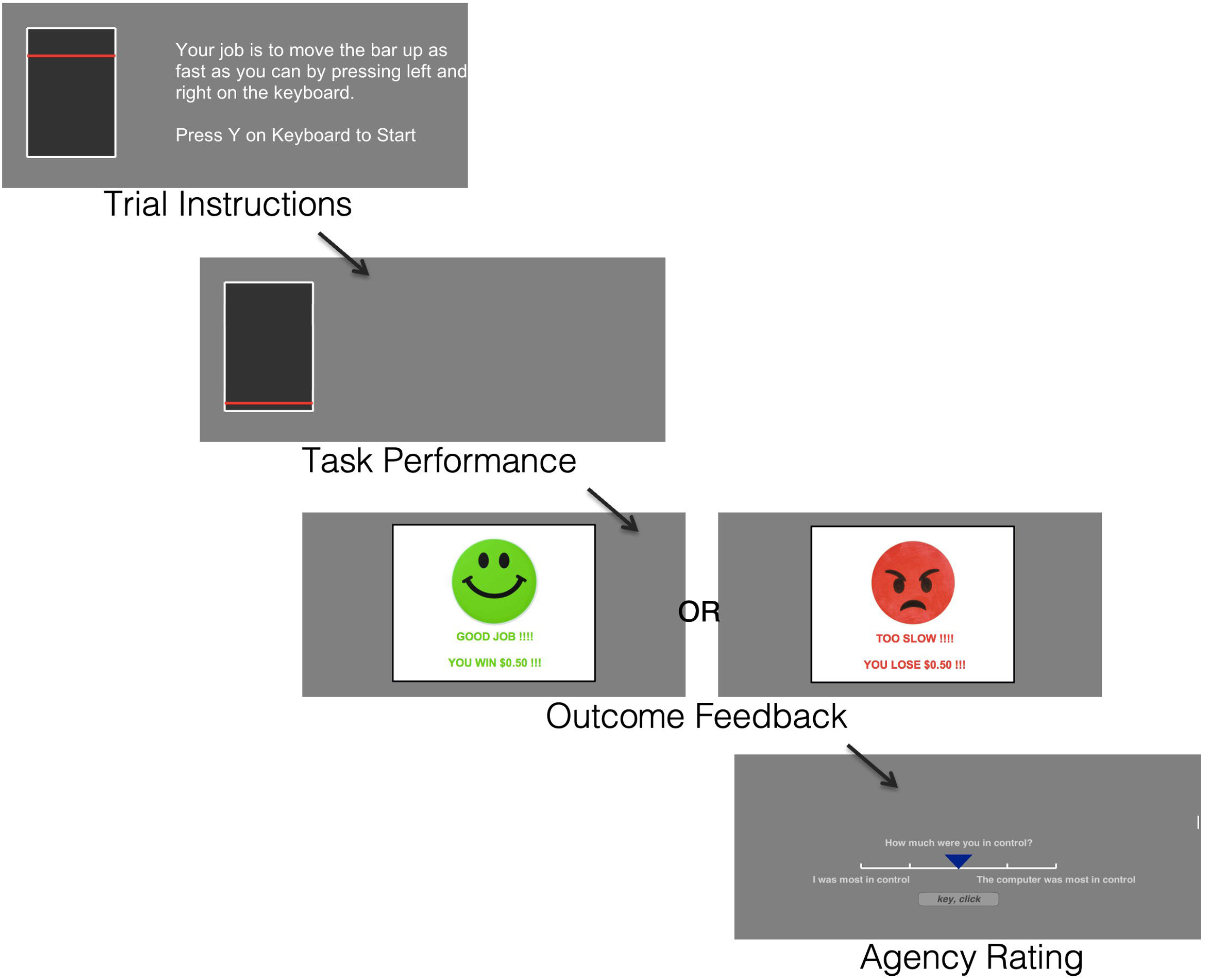
Self-Agency Task (SAT) schematic. On each trial, subjects perform a simple button press effort task for monetary reward. Subjects then gain positive-win or negative-loss feedback based on their reaction time relative to their own baseline reaction time. Following feedback, subjects are instructed to rate the level of agency for that trial on a 5-point Likert scale (1=self, 5=computer).

Subjects were instructed that during the task there may be computer interference as they perform the simple effort task. During the task, there were 3 conditions of computer interference to modulate experience of agency, which was not known to the subject. The *Self* condition includes 18 trials with no computer interference, whereby each pair of button presses (i.e. left + right) moved the bar by 1 position, thus 4 button presses moves the bar up by 2 positions, etc. The *Computer* condition includes 18 trials with maximum computer interference whereby button presses did not directly equate to incremental bar movement. Here, for each bar position, a randomly determined number of button presses (between 1-6) was required to move the bar by 1 position such that there was a dissociation between button presses and visual feedback of bar movement. The *Ambiguous* condition included 72 trials with a mix of trials that include mild computer interference (between 1-3 button presses required to move the bar by 1 position). At the end of each trial, subjects rated who was more “in control” during that trial on a 5-point Likert scale where 1=self, 5=computer (Figure 1). The majority of trials were *Ambiguous* as performance during this condition is the main behavioral measure of interest. The *Self* and *Computer* condition trials were included firstly as contrast trials to objectively indicate the varying levels of agency and secondly to allow measurement of overall task accuracy and understanding. Subjects undertook training on the task in a self-paced manner before starting the task, including instruction and experience of the maximum computer interference i.e., the “computer” condition. Subjects received bonus monetary payment depending on performance.

The online version of the SAT task was adapted for online testing, identical to the in-person version. The task was created with *PsychoPy3* and hosted on their online platform, Pavlovia (https://pavlovia.org/).

### Statistics

In order to determine the impact of outcome valence on self-agency, agency rating following positive-win outcomes and negative-loss outcomes (when computer interference was kept constant in *Ambiguous* trials) were subjected to paired-samples t-test for all subjects. Second, to examine group differences in self-agency, two-tailed independent samples t-test was conducted to determine differences in self-agency between HC and DA. Finally, a mixed-effects linear regression was conducted to identify how all variables predicted self-agency ratings, including the effects of valenced outcome feedback (i.e., win/loss), condition (i.e., self/computer/ambiguous), and group (i.e., HC/DA) on agency rating (rating ∼ 1 + group + agency + feedback + group:agency + group:feedback + (1 + agency + feedback | subject)). A separate mixed-effects linear regression was also conducted using separate groups of HC, MDD (primary) and GAD (primary) on an exploratory basis. The regression analysis was repeated with the online sample to evaluate how feedback and agency predicted ratings (rating ∼ agency + feedback + (1 + agency | subject) + (1 + feedback | subject). A mixed-effects linear model was used to account for individual differences. The regressions were carried out using the *fitlme* function in Matlab.

## Results

### Experiment 1: In Person Participants

A total of 52 HC subjects performed the task. Two HC subjects did not fully complete the task, leaving 50 HC subjects for analysis (age=39.4 ±9.9, 21 female). Twenty patients with DA disorders (age=33.4 ±8.8, 13 female) completed the task including 7 MDD primary, 13 GAD primary, with the majority of DA subjects experiencing significant comorbidity (see Table 1 for demographics).

**Table 1.** Participant characteristics for patients with depression and anxiety (DA) and healthy controls (HC). Anhedonia is measured with the temporal experience of pleasure scale (TEPS).

|  | Healthy Controls | Depression and Anxiety |
| --- | --- | --- |
| N | 50 | 20 |
| Age (mean ± SD) | 39.4 ± 9.93 | 33.35 ± 8.89 |
| Females (frequency, %) | 21, 42% | 15, 75% |
| Hispanic ethnicity (frequency, %) | 7, 14% | 4, 20% |
| Employed, at least part-time (frequency, %) | 30, 60% | 7, 35% |
| Some college (frequency, %) | 22, 90% | 17, 85% |
| Married (frequency, %) | 13, 26% | 4, 20% |
| N | 32 | 17 |
| Anhedonia (anticipatory) | 41.75 ± 8.04 | 40.18 ± 7.67 |
| Anhedonia (consummatory) | 37.34 ± 6.99 | 38.18 ± 8.88 |

#### Self-Agency Task

All subjects demonstrated higher self-agency after receiving positive-win feedback, compared to negative-loss feedback during the *Ambiguous* trials when the level of computer inference was kept constant (Figure 2, T=-6.25, p=1.5×10^−8^). This remained the case for the HC group (T=-5.04, p=0.000007) and the DA group (T=-4.07, p=0.001), separately (Figure 2).

**Figure 2.**
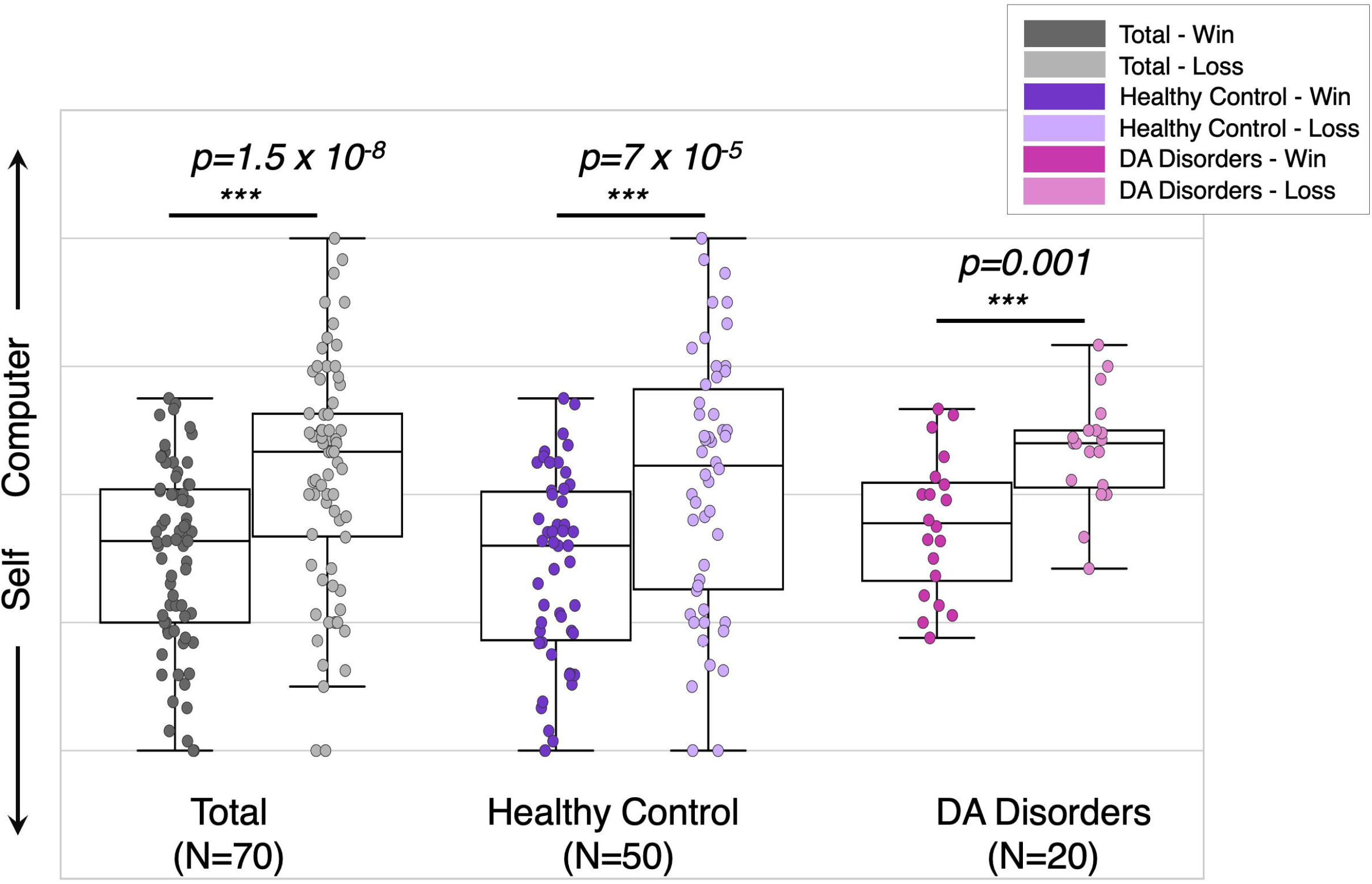
Higher positive valence-dependent self-agency across all groups. Agency ratings plotted following positive-win (darker colors) and negative-loss (lighter colors) feedback during Ambiguous trials for all subjects (N=70), healthy control subjects only (N=50) and subjects with depression and anxiety (DA) disorders (N=20). All subjects demonstrated higher self-agency following positive-win outcomes compared to negative-loss outcomes.

Higher self-agency following the positive-win feedback was associated with higher age across all subjects (R=-0.308, p=0.006) and for the HC group (R=-0.314, p=0.045) and the DA group (R=-0.470, p=0.037), separately (Figure 3). There was no relationship between age and self-agency following negative-lose feedback (p’s>0.3), indicating a specificity of this bias in the positive domain. There were no differences within any groups or across groups based on sex (p’s>0.5).

**Figure 3.**
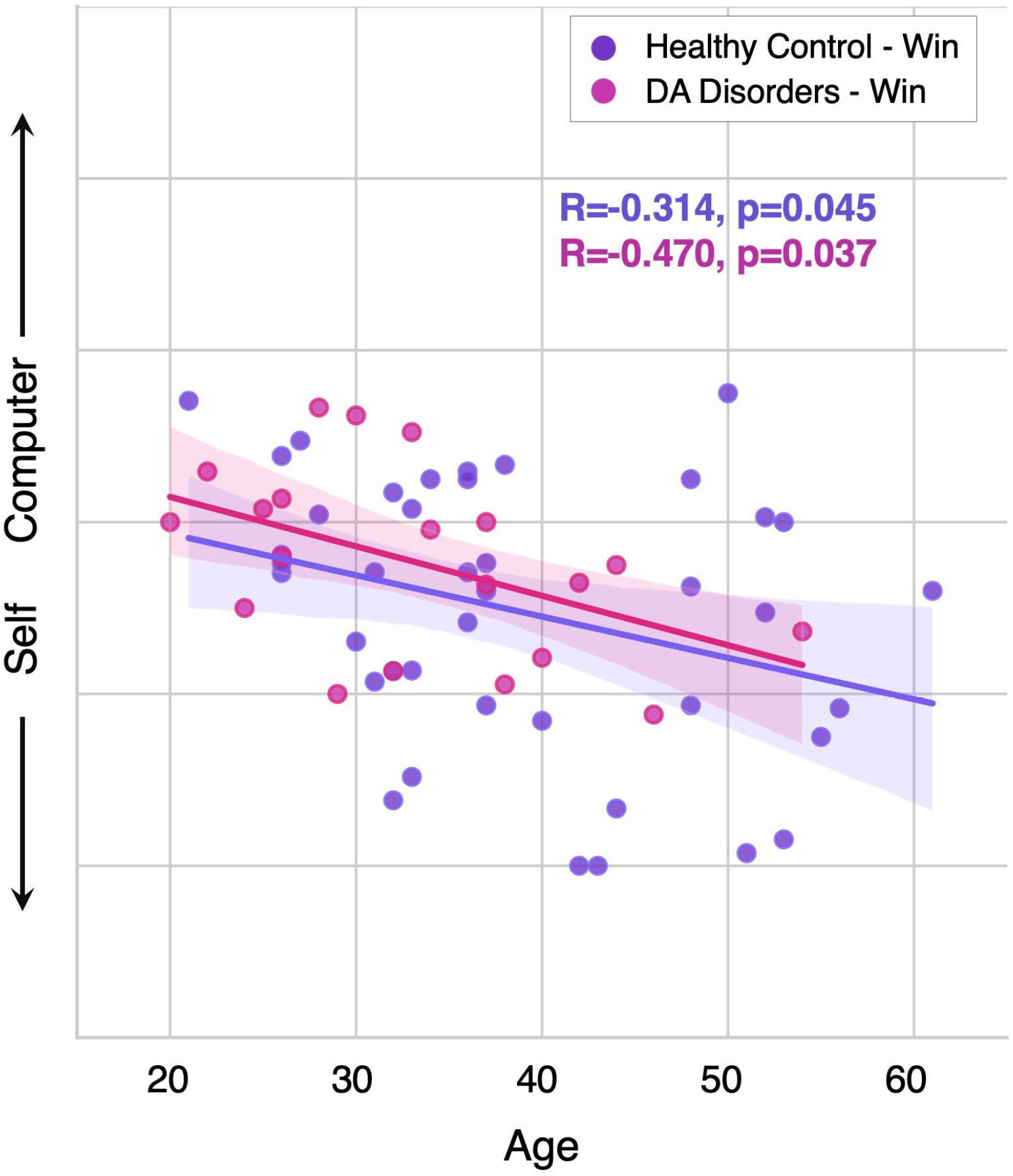
Higher positive valence-dependent self-agency increases with age across all groups. Agency ratings plotted following positive-win feedback against age in healthy control subjects (N=50, purple) and subjects with depression and anxiety (DA) disorders (N=20, pink).

Patients with DA disorders reported lower self-agency following positive-win feedback compared to healthy controls (T=-2.171, p=0.035) (Figure 4a), with no difference following negative-loss feedback (T=-1.475, p=0.145).

**Figure 4.**
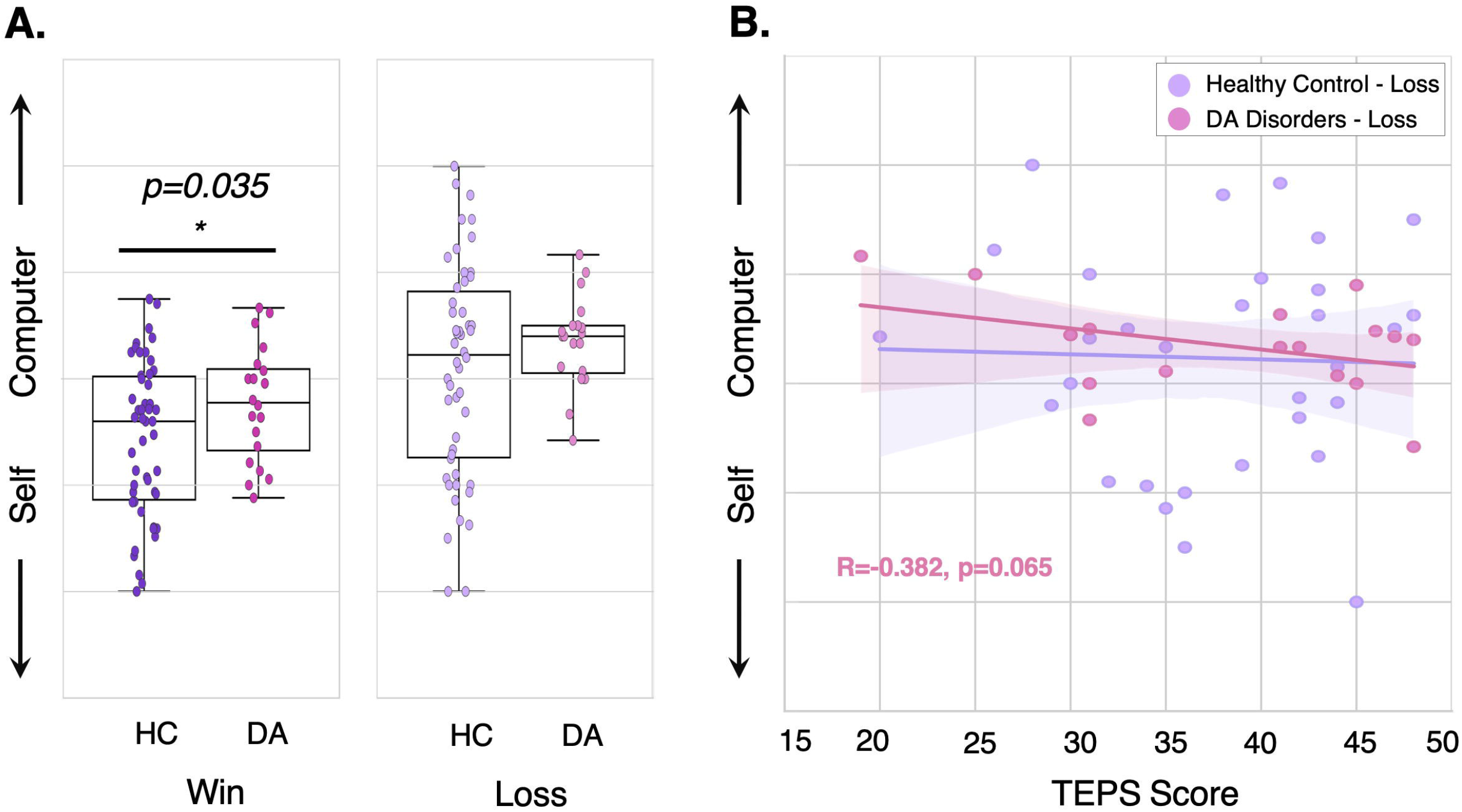
Lower negative valence-dependent self-agency in depression and anxiety disorders and relationships with symptoms. A, Agency ratings plotted following positive-win and negative-loss feedback during Ambiguous trials, plotted separately for healthy control subjects (N=50) and subjects with depression and anxiety (DA) disorders (N=20) (data is recapitulated from Figure 1). B, Agency ratings plotted following negative-loss feedback against Temporal Experience of Please Scale (TEPS) consummatory score for both groups. Higher TEPS scores represent higher pleasure ratings, thus being inversely related to anhedonia.

#### Mixed-effects Linear Regression Model

A mixed-effects linear regression was conducted to examine all effects, including group (HC/DA), agency condition (*Self, Computer, Ambiguous*), and outcome feedback (positive-win, negative-loss) on agency rating.

First, there was a significant effect of agency condition whereby self-agency was higher in the *Self* group (the reference group in the regression) than the *Ambiguous* condition (*β* = 0.40, *p* < 0.001) or the *Computer* condition (*β* = 1.16, *p* < 0.001), meaning subjects understood the task and performed accurately. Second, there was a significant effect of feedback valence (*β* = -0.66, *p* < 0.001) whereby there was higher self-agency following positive-win compared to negative-loss feedback, corroborating the primary test above. Third, there was no significant main effect of group (*β* = 0.01, *p* = 0.97), but there was a significant interaction between group and feedback (*β* = 0.27, *p* < 0.05), whereby DA subjects had lower self-agency following positive-win feedback compared to the HC group.

The second exploratory regression model that included separate denotations of MDD-primary and GAD-primary groupings produced largely similar results and suggested that both MDD-primary subjects (*β* = 0.38, *p* < 0.05) and GAD-primary subjects (*β* = 0.23, *p* = 0.06) had lower self-agency following positive-win feedback compared to HC. See Supplementary Tables 1-2 for all results from the mixed-effects regression models.

#### Dimensional symptom domains

Relationships between self-agency and anhedonia (TEPS) were examined in an exploratory manner. There were no significant relationships between self-agency and dimensional measures of anhedonia in HC (Figure 4b). In DA, lower self-agency following negative-loss feedback showed a trend towards correlation with worse self-reported symptoms of anhedonia (TEPS-consummatory, R=-0.382, p=0.065) (Figure 4b).

### Experiment 2: Online Participants

A total of 254 participants performed the task, of which, one participant did not fully complete the task. After the preliminary quality check, seven participants were excluded, leaving a sample size of 246 for analysis (age=32.83 ±6.54, 127 female). See Table 2 for subject characteristics.

**Table 2.**
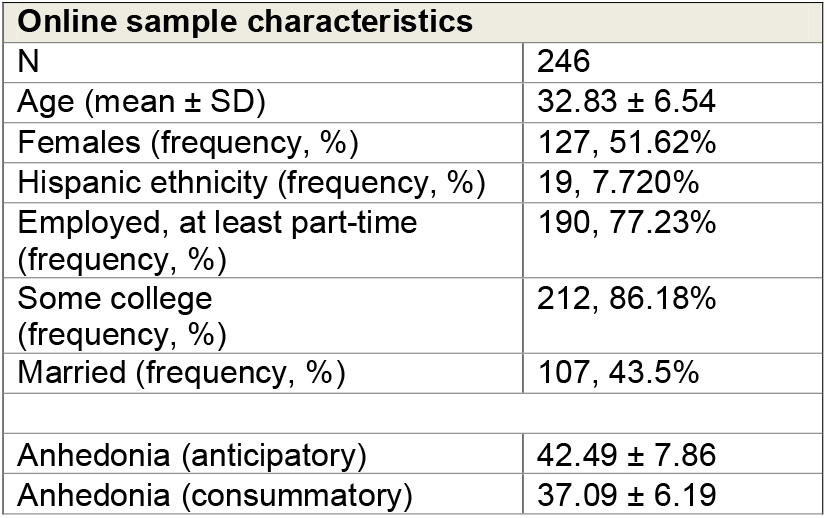
Participant characteristics for participants from the online study. Anhedonia is measured with the Temporal experience of pleasure scale (TEPS).

| Online sample characteristics |  |
| --- | --- |
| N | 246 |
| Age (mean ± SD) | 32.83 ± 6.54 |
| Females (frequency, %) | 127, 51.62% |
| Hispanic ethnicity (frequency, %) | 19, 7.720% |
| Employed, at least part-time (frequency, %) | 190, 77.23% |
| Some college (frequency, %) | 212, 86.18% |
| Married (frequency, %) | 107, 43.5% |
| Anhedonia (anticipatory) | 42.49 ± 7.86 |
| Anhedonia (consummatory) | 37.09 ± 6.19 |

#### Self Agency Task

Similar to the in-person findings, all subjects demonstrated higher self-agency after receiving positive-win feedback, compared to negative-loss feedback during the *Ambiguous* trials when the level of computer inference was kept constant (Figure 5A, T= 15.07, p= 8.57×10^−34^).

**Figure 5.**
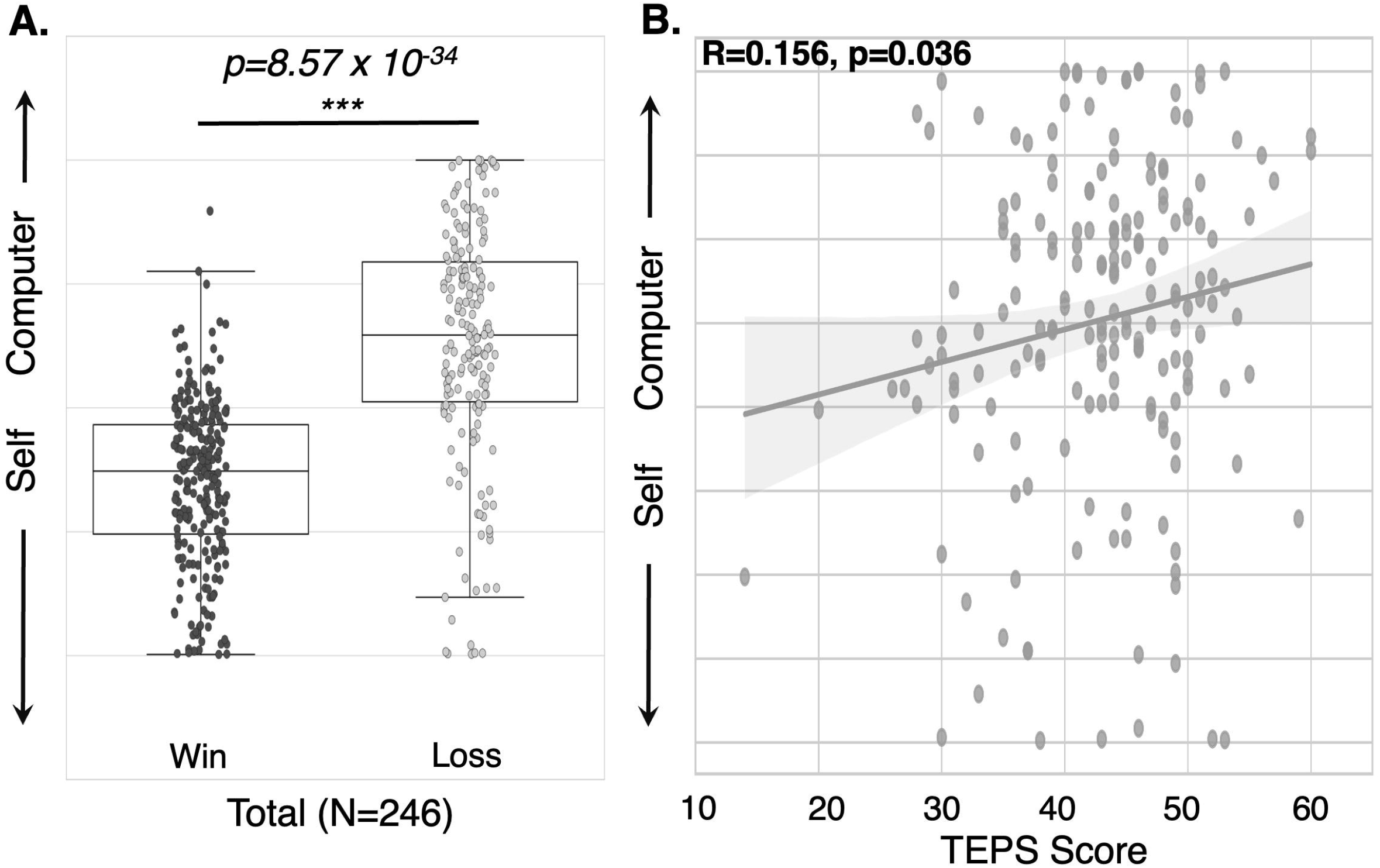
Higher positive valence-dependent agency replicated in online sample. A, Agency ratings plotted following positive-win and negative-loss feedback during Ambiguous trials for all subjects in the online session (N=246). B, Agency ratings plotted following negative-loss feedback during Ambiguous trials against Temporal Experience of Please Scale (TEPS) anticipatory sub-score for the online sample (N=246). Higher TEPS scores represent higher pleasure ratings, thus being inversely related to anhedonia.

Higher self-agency following the positive-win feedback showed a negative relationship, with higher age across all subjects, which did not reach significance (R=-0.1, p=0.135). The correlation was likely not significant due to the restricted age range of this sample (ages 18-45). Again, there was no relationship between age and self-agency following negative-lose feedback, and no differences based on sex (p’s>0.5).

#### Mixed-effects Linear Regression Model

A mixed-effects linear regression was conducted to examine all effects, including agency condition (*Self, Computer, Ambiguous*), and outcome feedback (positive-win, negative-loss) on agency rating. First, we replicated the significant effect of agency condition (β=0.39, p=6.13×10^−54^) whereby self-agency was higher in the *Self* group (the reference group in the regression) than the *Ambiguous* condition (T= -659, p= 1.16×10^−10^) and the *Computer* condition (T= -11.28, p= 2.24×10^−26^), meaning subjects understood the task and performed accurately.

Second, there was also a significant effect of feedback valence (β = 1.10, *p =* 1.78 ×10^−64^) whereby there was higher self-agency following positive-win compared to negative-loss feedback (T= - 14.05, p= 9.82×10^−36^), corroborating the primary test above. See Supplementary Table 3 for the results from the mixed-effects regression model.

#### Dimensional symptom domains

Relationships between self-agency and anhedonia were again examined in an exploratory manner across all online subjects. Similar to the findings in Experiment 1, lower self-agency following negative-loss feedback was associated with worse symptoms of anhedonia (TEPS-anticipatory) in the online sample (R=0.156, p=0.036, Figure 5B).

## Discussion

This paper showcases an objective study of self-agency in a dynamic environment in healthy individuals and subjects with mood and anxiety disorders. Since self-agency has been with syndromes related to anhedonia in depression (Soral et al., 2021; Thompson et al., 2013; Yoshie & Haggard, 2013) and high risk for problematic anxiety (Chorpita & Barlow, 1998), this study focused on the symptom domain of anhedonia and its relationship with sense of self-agency in the context of positive and negative environments. The task was also repeated in an online sample, representative of a general population to capture variations in anhedonia reporting and relationships with agency.

Across all subjects in both experiments, there was higher self-agency in a positive rewarding environmental and lower self-agency in a negative environment. This positive valence-dependent agency generally increased with increasing age. Subjects with mood and anxiety disorders showed lower negative valence-dependent agency compared to healthy controls, with no difference during the negative context. Higher self-agency in a negative environment was also associated with worse self-reported anhedonia symptoms across both experiments. Together these results indicate a novel belief bias that presents across all subjects and suggests a potential trans-diagnostic disturbance related to reduction in self-agency.

The current results corroborate literature on self-agency bias in healthy subjects that increases with age (Hovenkamp-Hermelink et al., 2019). However, this extends upon previous work by utilizing an objective cognitive task that directly separates self-agency in a positive context compared to a negative context, highlighting that only the positive valence-dependent agency increases with age.

Reduced negative-valence dependent agency in subjects with DA disorders was expected given previous literature linking self-reported self-agency and self-reported depression (Abdolmanafi et al., 2011) and anxiety (Hovenkamp-Hermelink et al., 2019) symptoms. However, the current findings suggest that in this sample of patients, reduced self-bias or a more external locus of control exists more so in a positive environment only, rather than a negative environment. This is surprising given a wealth of literature highlighting negative biases in depression and anxiety disorders. Selective memory recall for negative information (Hovenkamp-Hermelink et al., 2019) and negative bias related to perception of facial expressions (Raes et al., 2006) is higher in depressed cohorts. Similarly, attentional bias towards negative information (Mogg & Bradley, 2005) and negative interpretation bias of ambiguous information (Huppert et al., 2003) is well established in anxiety disorders. However, while negative attentional bias is inconsistently reported in depression, this bias seems to increase for self-relevant negative information (Mogg et al., 2006; Segal et al., 1995), whereby depressed subjects relate negative information to self more than control subjects. This suggests that negative self-biases in depression may relate to negative self-evaluation, rather than in terms of negative environmental outcomes of ones’ actions. While there were no group differences in negative self-bias in these samples, higher negative self-bias was associated with anhedonia in patients and in a larger population-based cohort. These results suggest an interesting avenue for exploring possible relationships between anhedonia symptoms related to lower negative-valence dependent agency. However, these findings must be replicated in another study before confirming specific links between anhedonia and negative valence-dependent self-bias.

Finally, while mood and anxiety disorders and symptoms are highly comorbid (J. D. Brown et al., 2001; T. A. Brown et al., 2001), further studies in larger sample sizes must disentangle the relative contributions of positive and negative valence-dependent self-agency to depression, anhedonia, stress and anxiety –related symptoms across and within disorder groups. Moving away from categorical diagnoses of disorders, it will be important to dissect how specific symptom domains, such as rumination or threat-reactivity relate to underlying disturbances in self-agency across groups. If these findings are replicated, an important future avenue of research would be to examine how agency belief-updating can be modulated using self-agency related feedback or training, in order to normalize aberrant self-agency biases related to symptoms across patient groups.

## Data Availability

All data produced in the present study are available upon reasonable request to the authors

## Data Availability

The datasets generated during and/or analyzed during the current study are available from the corresponding author upon request.

