## Supplementary Data for "Valence-dependent self-agency is disturbed in depression and anxiety"

| Independent | Predictor | Estimate | SE | t-value | P |
| --- | --- | --- | --- | --- | --- |
| Rating | **Intercept** | 2.65 | 0.13 | 20.76 | <0.001 |
|  | Group - DA | 0.01 | 0.18 | 0.04 | 0.972 |
|  | **Agency (*Ambiguous*)** | 0.40 | 0.09 | 4.35 | 0.000 |
|  | **Agency (*Computer*)** | 1.16 | 0.15 | 7.82 | 0.000 |
|  | **Feedback (*Win*)** | -0.66 | 0.11 | -6.04 | 0.000 |
|  | Group (*DA*) x Agency (*Ambiguous*) | 0.22 | 0.16 | 1.36 | 0.175 |
|  | Group (*DA*) x Agency (*Computer*) | -0.05 | 0.23 | -0.20 | 0.842 |
|  | Group (*DA*) x Feedback (*Win*) | 0.27 | 0.11 | 2.46 | 0.014 |

Supplementary Table 1. Results of the Linear Mixed Effects Model to test agency and feedback as predictors of rating (sense of agency) for patients with Depression and Anxiety Disorder (DA).

| **Independent** | **Predictor** | **Estimate** | **SE** | **t-value** | **P** |
| --- | --- | --- | --- | --- | --- |
| Rating | **Intercept** | 2.65 | 0.13 | 20.72 | <0.001 |
|  | Group (*GAD*) | 0.01 | 0.21 | 0.05 | 0.96 |
|  | Group (*MDD)* | 0 | 0.25 | 0.02 | 0.985 |
|  | **Agency (*Ambiguous*)** | 0.4 | 0.09 | 4.4 | <0.001 |
|  | **Agency (*Computer*)** | 1.16 | 0.15 | 7.81 | <0.001 |
|  | **Feedback (*Win*)** | -0.66 | 0.11 | -6.09 | <0.001 |
|  | Group (*GAD*) x Agency (*Ambiguous*) | 0.13 | 0.19 | 0.72 | 0.473 |
|  | Group (*MDD*) x Agency (*Ambiguous*) | 0.38 | 0.23 | 1.66 | 0.097 |
|  | Group (*GAD*) x Agency (*Computer*) | -0.09 | 0.26 | -0.33 | 0.738 |
|  | Group (*MDD*) x Agency (*Computer*) | 0.01 | 0.33 | 0.04 | 0.969 |
|  | Group (*GAD*) x Feedback (*Win*) | 0.23 | 0.12 | 1.91 | 0.056 |
|  | **Group (*MDD*) x Feedback (*Win*)** | 0.38 | 0.16 | 2.39 | 0.017 |

Supplementary Table 2. Results of the Linear Mixed Effects Model to test agency and feedback as predictors of rating (sense of agency) for patients with primary Generalized Anxiety Disorder (GAD) and Major Depressive Disorder (MDD), which compromised the Depression and Anxiety Disorder (DA) group.

| **Independent** | **Predictor** | **Estimate** | **SE** | **t-value** | **P** |
| --- | --- | --- | --- | --- | --- |
| Rating | **Intercept** | 2.0074 | 0.040489 | 49.579 | <0.001 |
|  | **Feedback** | 1.1045 | 0.064793 | 17.047 | 1.78E-64 |
|  | **Agency** | 0.39541 | 0.025456 | 15.533 | 6.13E-54 |

Supplementary Table 3. Results of the Linear Mixed Effects Model to test agency (self, ambiguous and computer) and feedback (positive-win and negative-loss) as predictors of rating (sense of agency) for the online sample.
